# Summary of the Effects of Exercise Therapy in Non-Communicable Diseases: Clinically Relevant Evidence from Meta-Analyses of Randomized Controlled Trials

**DOI:** 10.1101/2021.02.11.21251608

**Authors:** Urho M. Kujala

## Abstract

There is strong evidence that exercise therapy leads to better measured and self-reported physical fitness and function in patients with chronic diseases, along with improvements in body composition. The evidence for other health benefits has not been summarized as systematically across different chronic diseases. Given the volume of research that has appeared in the past decade, this review of meta-analyses of randomized controlled trials (RCTs) in patients with specific chronic diseases summarizes the evidence regarding exercise therapy for various outcomes to help practitioners in prescribing exercise therapy for their patients. Meta-analyses published before Jan 1, 2021, based on at least four RCTs, and investigating the effect of exercise therapy on the same outcome among patients with a specific chronic disease were screened. These meta-analyses show that, in addition to improvements in fitness and function, various cardio-metabolic risk factor levels are improved in most of the common cardio-metabolic diseases, pain is reduced in musculoskeletal diseases, mood (depression and anxiety) and health-related quality of life are improved in various disease categories, and disease-specific indicators of disease progression are improved for conditions such as type 2 diabetes, hypertension, coronary heart disease, heart failure, claudication, chronic obstructive pulmonary disease, rheumatoid arthritis, fibromyalgia, depression, anxiety, and schizophrenia. Physicians should consider prescribing exercise to their patients with chronic disease conditions to improve their physical fitness, mood, and health-related quality of life and to slow down the progression of disease. This improves a patient’s possibility to enjoy an active and independent life.

**Key points:**

- This study summarizes the data from meta-analyses of randomized controlled trials (RCTs) on the effects of exercise therapy in patients and discusses the clinical relevance of the findings.
- The findings include the following: various cardio-metabolic risk factor levels are improved in most of the common cardio-metabolic diseases, pain is reduced in musculoskeletal diseases, mood (depression and anxiety) and health-related quality of life are improved in various disease categories, and disease-specific indicators of disease progression are improved in conditions such as type 2 diabetes, hypertension, coronary heart disease, heart failure, claudication, chronic obstructive pulmonary disease, fibromyalgia, depression, anxiety, and schizophrenia.
- RCTs usually test the same exercise program for all trial participants. To tailor the exercise therapy intervention in real life, individually based factors, such as disease status, fitness level, personal exercising interests and possibilities, and the infrastructure making different exercise therapy modalities possible in the patient’s everyday environment should all be considered.

## Introduction

There is strong evidence from randomized controlled trials (RCTs) involving patients with chronic disease that exercise therapy leads to better measured and reported physical fitness and function [1]. In addition, exercise training leads to improvements in body composition, particularly controlling body weight and the reduction of visceral fat [2-5]. Epidemiological research shows an association between high physical activity and low risk of death, but on the basis of meta-analyses of high-quality RCTs, exercise therapy does not statistically significantly reduce all-cause mortality [6-8]. However, there remain many potential health benefits of exercise therapy in patients with chronic diseases that fall between the well-demonstrated effects of exercise on fitness and the inconclusive evidence on reducing all-cause mortality.

Genetic factors, many known and unknown confounding factors, and reverse causality may explain the associations between physical activity levels and documented health benefits in observational population follow-ups [7]. To use the resources of health care, strong evidence of those benefits in both theory and real life is needed. Thus, this review is based on clinical experience and meta-analyses of RCTs to summarize the strongest evidence regarding exercise therapy in the treatment of non-communicable diseases (NCDs) among adults. The outcomes reviewed are cardio-metabolic risk factor levels, pain, mood (depression and anxiety), disease-specific indicators of disease progression, and health-related quality of life (HRQoL). The overall goal is to summarize which benefits are based on strong interventional evidence when tailoring exercise therapy for patients. As large amounts of data have been reported during the past decade, this type of summary is important for health care and exercise sector practitioners and decision makers to know for which purposes exercise therapy should primarily be used and which exercise effects are most justified in being recommended to patients and motivate them to exercise.

## Methods

### Review and search strategy and selection criteria of included meta-analyses

To best serve readers, this review combines clinical, real-life experience with repeated semi-systematic literature searches. Formally, evidence from meta-analyses with at least 4 randomized controlled trials, a total of at least 150 participants, and investigating the effects of exercise therapy on the same outcome among patients with a specific chronic disease is summarized. For studies to be included in this review, the intervention and the control groups had to be primarily contrasted by physical exercise intervention (exercise therapy) and the participants of the trials had to be primarily adults. The protocol is designed for this purpose and does not exactly match earlier published protocol guidelines; it was conducted by only one author.

Data for this review were identified by various yearly literature searches between 2000 and Jan 1, 2021, complemented by searches of PubMed and the Cochrane Database of Systematic Reviews and references in relevant articles through Jan 1, 2021 in early 2021. The design of this review partly follows the design of previous reviews by the same author focusing on the effects of exercise therapy in patients with chronic diseases [9, 10]. However, in this updated version, the findings are classified according to outcomes instead of diseases to evaluate whether similar results are achieved in patients with different chronic diseases, and follow-up in the literature is extended by 12 years compared to the previous review [10]. Large amounts of new data (and most of the data presented here) have appeared in the past decade (see Supplementary file 1).

The searches and this review systematically cover the following NCDs: metabolic syndrome, diabetes, hypertension, coronary heart disease, heart failure, claudication, stroke, asthma, chronic obstructive pulmonary disease (COPD), osteoarthritis, rheumatoid arthritis, low back pain, neck pain, fibromyalgia, osteoporosis, depression, anxiety, schizophrenia, dementia, Parkinson’s disease, multiple sclerosis, cancers, and chronic fatigue syndrome, along with individuals living with human immunodeficiency virus (HIV). For search terms of the latest confirmatory searches, see Supplementary file 1. Obesity alone is not included as a disease in this review. The final searches were limited to full-text availability of items written in English. Results from initial follow-ups, usually including the most comprehensive data from the RCTs included in the meta-analyses, are primarily reported. No criteria as to length of follow-ups were established.

Only results with statistically significant effects of exercise therapy compared to controls are included. Generally, the newest meta-analyses are included; older meta-analyses with the same disease and same outcome are not included if they report the same results as the newer ones. In addition, if there was a positive finding in an earlier meta-analysis that was not confirmed by a later meta-analysis with a larger number of trials or higher-quality studies, the meta-analysis results are not included. Due to the large number of outcome variables, only the most clinically important outcomes (based on the author’s view as informed by consulting other specialists and clinical benefits for the patient and/or health care system) are included.

The primary interest was to see whether land-based exercise therapy causing physiological loading to the whole body provides benefits; the focus was not on particular types of training. Very specific training modalities such as training respiratory muscles are not included. However, differences in the effects of different types of exercise, such as aerobic versus strength training are discussed when clinically relevant. So-called mind-body exercises, which combine mental and physical exercises (Pilates, tai chi, yoga, and health qigong, and games) are not included in this review; although they may be effective treatment methods in many chronic diseases, their effects may not be due to physical exercise per se.

Meta-analyses do not always report whether the collected results are based on “intention-to-treat” or “per protocol” results of included RCTs. When the results are based on the analysis of an intention to treat, those results are included in this review.

In clinical work, prescribing exercise to formerly inactive patients is often combined with recommendations regarding other health habits such as diet changes, reducing or eliminating smoking or the use of alcohol, but this review concentrates on the independent effects of exercise therapy. Meta-analyses of studies in which other types of interventions have clearly been added to exercise intervention but do not apply to the control group are not included in this review.

### Summary of findings on the effects of exercise therapy

In the findings (see Tables 1-5), only meta-analyses providing statistically significant effects sizes on the effects of exercise therapy on clinically significant outcomes are included.

**Table 1.** Effects of exercise therapy on cardio-metabolic risk factors, based on selected meta-analyses of randomized controlled trials in the treatment of patients with specific diseases.

| Study | Disease | Outcome measure | N trials<br>(N participants) | Effect size of exercise compared to controls, pooled statistics (95% CI)<sup>a</sup> |
| --- | --- | --- | --- | --- |
| Wewege et al. [4]<sup>b</sup> | Metabolic syndrome | Waist circumference | 13 (504) | Mean difference -3.44 cm (-4.81, -2.07) |
| Wewege et al. [4]<sup>b</sup> | Metabolic syndrome | Fasting blood glucose | 11 (471) | Mean difference -0.15 mmol/L (-0.29, -0.02) |
| Wewege et al. [4]<sup>b</sup> | Metabolic syndrome | HDL cholesterol | 9 (427) | Mean difference 0.05 mmol/L (0.01, 0.08) |
| Wewege et al. [4]<sup>b</sup> | Metabolic syndrome | Triglycerides | 11 (471) | Men difference -0.29 mmol/L (-0.43, -0.14) |
| Yousefabadi et al. [11] | Metabolic syndrome | C-reactive protein (CRP) | 29 (1096) | Weighted mean difference -0.52 mg/L (-0.79, -0.25) |
| Yousefabadi et al. [11] | Metabolic syndrome | Tumor necrosis factor-α (TNF-α) | 23 (696) | Weighted mean difference -1.21 pg/mL (-1.77, -0.66) |
| Wewege et al. [4]<sup>b</sup> | Metabolic syndrome | Diastolic blood pressure | 10 (336) | Mean difference -0.59 mmHg (-2.86, -0.32) |
| Umpierre et al. [12] | Type 2 diabetes | Glycated hemoglobin (HbA<sub>1c</sub>) percent | 23 (1533) | Weighted mean difference -0.67% (-0.84%, -0.49%) |
| Kelley & Kelley [13]<sup>b</sup> | Type 2 diabetes | LDL cholesterol | 4 (156) | Weighted mean difference -6.4 mg/dL (-11.8, -1.1) |
| Chen et al. [14] | Type 2 diabetes | CRP | 18 (996) | Weighted mean difference -0.79 mg/L (-1.26, -0.33) |
| Chen et al. [14] | Type 2 diabetes | TNF-α | 10 (350) | Weighted mean difference -2.33 μg/mL (-3.39, -1.27) |
| Lee et al. [15] | Type 2 diabetes | Brachial artery flow mediated dilatation | 13 (306) | Hedges' g 0.41 (0.21, 0.62) |
| Nolan et al. [15] | Coronary heart disease | Heart rate variability | 16 (631) | Standardized mean difference 0.36 (0.18, 0.55) |
| Taylor et al. [17] | Coronary heart disease | Total cholesterol | 17 (1903) | Weighted mean difference -0.37 mmol/L (-0.63, -0.11) |
| Kelley et al. [18]<sup>b</sup> | Cardiovascular disease | HDL cholesterol | 6 (637) | Weighted mean difference 3.7 mg/dL (1.2, 6.1) |
| Taylor et al. [17] | Coronary heart disease | Triglycerides | 13 (1442) | Weighted mean difference -0.23 mmol/L (-0.39, -0.07) |
| Kelley et al. [18]<sup>b</sup> | Cardiovascular disease | Triglycerides | 9 (1172) | Weighted mean difference -19.3 mg/dL (-30.1, -8.5) |
| Zhang & Chang [19] | Coronary heart disease with percutaneous coronary intervention | Left ventricle ejection fraction | 5 (735) | Weighted mean difference 2.82% (1.50, 4.14) |
| Brouwer et al. [20]<sup>b</sup> | Stroke | Systolic blood pressure | 6 (377) | Mean difference -3.59 mmHg (-6.14, -1.05) |
| Brouwer et al. [20]<sup>b</sup> | Stroke | Fasting glucose | 4 (184) | Mean difference -0.12 mmol/L (-0.23, -0.02) |
| Heiwe et al. [21] | Chronic kidney disease | Systolic blood pressure | 9 (347) | Mean difference -6.08 mmHg (-10.12, -2.15) |
| Zang et al. [22] | Non-dialysis chronic kidney disease | Systolic blood pressure | 9 (463) | Mean difference -5.61 mmHg (-8.99, -2.23) |
| Heiwe et al. [21] | Chronic kidney disease | Diastolic blood pressure | 11 (419) | Mean difference -2.32 mmHg (-4.05, -0.59) |
| Zang et al. [22] | Non-dialysis chronic kidney disease | Diastolic blood pressure | 8 (399) | Mean difference -2.87 mmHg (-3.65, -2.08) |

<sup>a</sup> All estimates reported in this table favor exercise groups; effect sizes are as reported by the authors of original meta-analyses.
<sup>b</sup> The meta-analysis includes only aerobic training trials.

### Effects on cardio-metabolic disease risk factors

As expected, changes in cardio-metabolic risk factor levels have been studied most in patients with metabolic syndrome, type 2 diabetes (T2D), and cardio-vascular diseases; exercise-induced improvements in the risk factors have been documented, according to the meta-analyses (Table 1) [4, 11-22]. The findings also include effects on several measures: improvement in blood lipids; reductions in inflammatory parameters, fasting glucose, glycated hemoglobin (HbA_1c_) percent, and systolic blood pressure; and improvements in flow-mediated dilatation of brachial artery, heart rate variability, and left ventricular ejection fraction (Table 1). In chronic kidney disease, exercise reduces blood pressure levels [21, 22]. Although following the selection criteria of this review these findings have been documented by meta-analyses only in specific diseases (Table 1), many of these trends also apply to other chronic diseases, according to individual RCTs.

Interestingly, a meta-regression analysis of 26 RCTs on exercise training in T2D patients showed that exercise frequency in supervised aerobic training and weekly volume of resistance exercise in supervised combined training are associated with a reduction in HbA_1c_ percent [23]. A recent meta-analysis by Pan et al. [24] confirmed that both supervised aerobic and resistance exercises reduced HbA_1c_ compared to no exercise, and both also reduced triacylglycerol (triglyceride) levels and increased high-density lipoprotein (HDL) cholesterol levels. So, at least for HbA_1c_ levels, resistance training reducing skeletal muscle glucose intolerance appears to be a good training modality apart from aerobic training.

### Effects on pain

The beneficial effects of exercise therapy on self-reported pain have been shown in chronic musculoskeletal disorders including osteoarthritis, rheumatoid arthritis, nonspecific chronic low-back pain, neck pain, and fibromyalgia (Table 2) [25-36]. Tanaka et al. [37] reported that non-weight-bearing strengthening exercises are more effective than weight-bearing aerobic exercises on short-term pain relief in osteoarthritis. Byström et al. [38] reported that, in patients with chronic low back pain, motor control exercises appear to be more effective than general exercises, while Wang et al. [39] reported that, in patients with chronic low back pain, core stability exercises appear to be more effective than general exercise with regard to pain reduction. However, differentiating between motor control, core stability, and other exercises is not a simple task.

**Table 2.** Effects of exercise therapy on pain, based on selected meta-analyses of randomized controlled trials in the treatment of patients with specific diseases.

| Study | Disease | Outcome measure | N trials (N participants) | Effect size of exercise compared to controls, pooled statistics (95% CI)<sup>a</sup> |
| --- | --- | --- | --- | --- |
| Fransen et al. [25] | Knee osteoarthritis | Pain | 44 (3537) | Standardized mean difference -0.49 (-0.59, -0.39) |
| Fransen et al. [26] | Hip osteoarthritis | Pain | 9 (549) | Standardized mean difference -0.38 (-0.55, -0.20) |
| Goh et al. [27] | Hip or knee osteoarthritis | Pain | 69 (5272) | Standardized mean difference -0.56 (-0.68, -0.44) |
| Baillet et al. [28]<sup>b</sup> | Rheumatoid arthritis | Pain | 6 (261) | Standardized mean difference -0.31 (-0.55, -0.06) |
| Hayden et al. [29] | Nonspecific chronic (>12 weeks) low back pain | Pain, visual analogue scale (scaled 0 to 100 points) | 8 (370) | Weighted mean difference -10.20 points (-19.09, -1.31) |
| Peek & Stevens [30] | Chronic low back pain | Pain | 45 (4462) | Standardized mean difference -0.32 (-0.44, -0.19) |
| Chen et al. [31] | Neck pain | Pain | 5 (605) | Standardized mean difference -0.59 (-0.29, -0.89) |
| Liang et al. [32] | Cervical radiculopathy | Pain | 9 (751) | Standardized mean difference -0.89 (-1.34, -0.44) |
| Busch et al. [33] | Fibromyalgia | Tender points | 6 (349) | Standardized mean difference -0.76 (-1.53, 0.01) |
| Sosa-Reina et al. [34] | Fibromyalgia | Pain | 9 (782) | Standardized mean difference -1.11 (-1.52, -0.71) |
| Bidonde et al. [35]<sup>b</sup> | Fibromyalgia | Pain | 6 (351) | Difference in absolute improvement 11% (4%, 18%) |
| Bidonde et al. [36] | Fibromyalgia | Pain | 15 (832) | Difference in absolute improvement 5% (1%, 9%) |

<sup>a</sup> All estimates reported in this table favor exercise groups; effect sizes are as reported by the authors of the original meta-analyses.
<sup>b</sup> The meta-analysis includes only aerobic training trials.

### Effects on mood (depression and anxiety)

Symptoms of depression and/or anxiety are reduced not only among patients with a clinical diagnosis of depression and/or anxiety but also in patients with other diseases, including diabetes, arthritis and other rheumatic diseases, Alzheimer’s disease, multiple sclerosis, and renal disease; cancer survivors and HIV-positive individuals also saw a reduction in such symptoms (Table 3) [40-48]. The evidence for the reduction of depression is strongest among breast cancer and lymphoma patients [44]. Herring et al. [49] studied the effect of exercise training on depression among patients with any chronic disease and, on the basis of 144 RCTs, the standardized mean difference (SMD) in the effect size for reduction of depression compared to control groups was 0.30 (95% CI, 0.25-0.36). Similarly, anxiety has been shown to decline among chronically ill patients as a result of exercise training (SMD compared to controls 0.29; 95% CI, 0.23-0.36) [50].

**Table 3.**
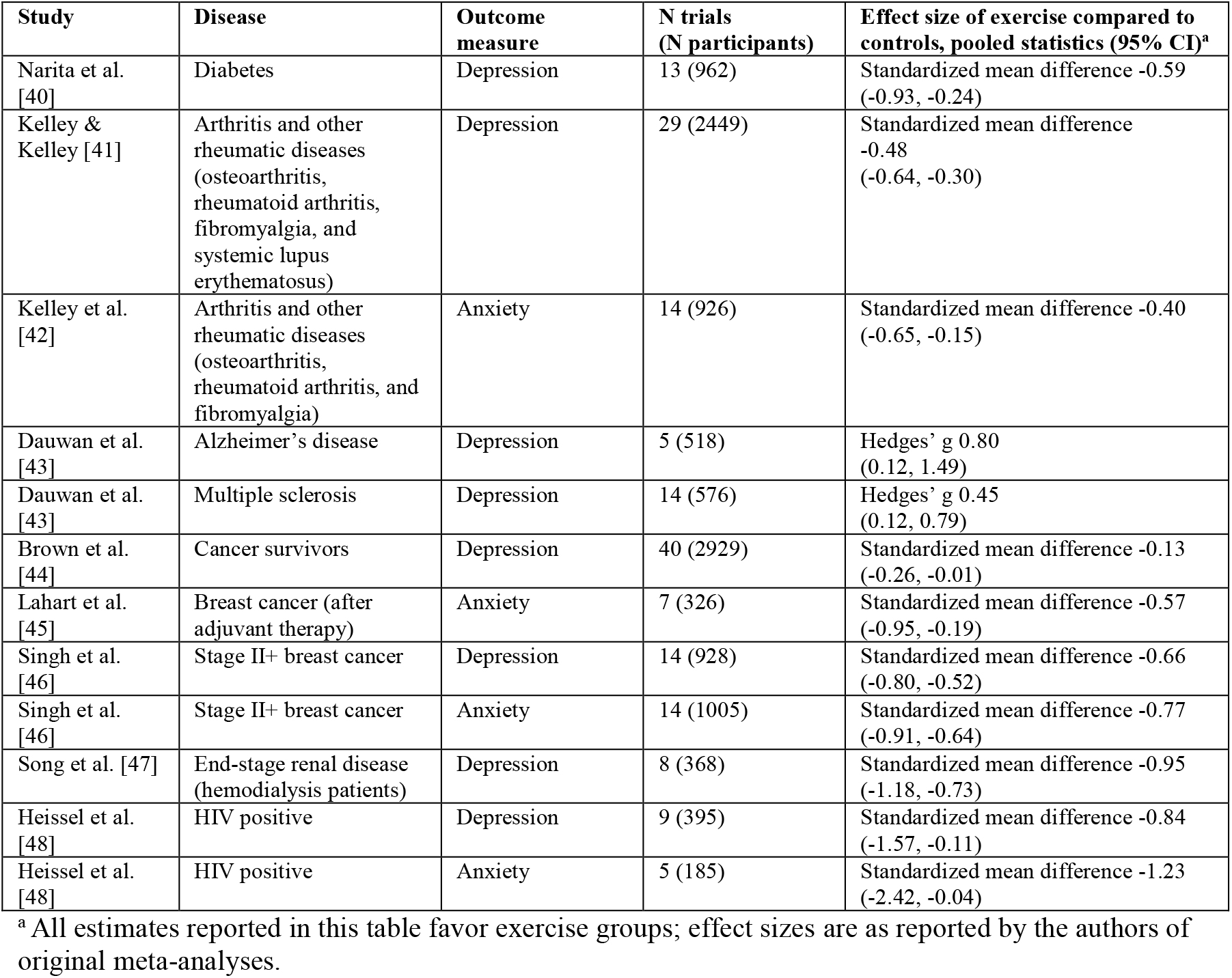
Effects of exercise therapy on mood (depression and anxiety), based on selected meta-analyses of randomized controlled trials in the treatment of patients with specific diseases.

### Effects on progression of chronic diseases

Theoretically, to have a real effect on disease progression and complications, a longer period of treatment is usually needed to have an effect on physical fitness or risk factor levels; however, there is a growing amount of data from RCT-based meta-analyses in which the main outcome is related to disease progression (Table 4) [8, 28, 32, 34, 51-67]. There is no single generally accepted criterion for following up on the progression of each chronic disease, but disease-specific clinical indicators such as findings from imaging studies, need for hospitalizations due to progression, or disease-specific scores of disease activity or stage can be used to characterize some aspects of disease severity or progression. The general HRQoL and disease-specific questionnaires that seek to assess disease status have overlapping features and cannot be distinctly categorized.

**Table 4.** Effects of exercise therapy on disease-specific indicators of disease severity/progression, based on selected meta-analyses of randomized controlled trials in the treatment of patients with specific diseases.

| Study | Disease | Outcome measure | N trials (N participants) | Effect size of exercise compared to controls, pooled statistics (95% CI)<sup>a</sup> |
| --- | --- | --- | --- | --- |
| Grace et al. [51]<sup>b</sup> | Type 2 diabetes | Glycated hemoglobin (HbA<sub>1c</sub>) percent | 20 (1347) | Mean difference -0.69% (-1.09, -0.36) |
| Grace et al. [51]<sup>b</sup> | Type 2 diabetes | Homeostatic model assessment of insulin resistance (HOMA-IR) | 7 (442) | Mean difference -1.02 (-1.77, -0.28) |
| Delevatti et al. [52]<sup>b</sup> | Type 2 diabetes | Glycated hemoglobin (HbA<sub>1c</sub>) percent | 26 (825) | Weighted mean difference -1.04 (-1.39, -0.69) |
| Cornelissen & Fagard [53]<sup>b</sup> | Hypertension | Systolic blood pressure | 30 (492) | Mean net change -6.9 mm Hg (-9.1, -4.6) |
| Cornelissen & Fagard [53]<sup>b</sup> | Hypertension | Diastolic blood pressure | 30 (492) | Mean net change -4.9 mm Hg (-6.5, -3.3) |
| Cao et al. [54]<sup>b</sup> | Hypertension | Systolic blood pressure | 13 (757) | Weighted mean difference -12.26 mmHg (-15.17, -9.34) |
| Cao et al. [54]<sup>b</sup> | Hypertension | Diastolic blood pressure | 13 (755) | Weighted mean difference -6.12 (-7.76, -4.48) |
| Heran et al. [55] | Coronary heart disease | Hospital admissions (6 to 12 mo follow-up) | 4 (463) | Risk ratio -0.69 (-0.93, -0.51) |
| Lawler et al. [56] | Myocardial infarction | Reinfarction | 18 (2550) | Pooled odds ratio 0.54 (0.38, 0.79) |
| Taylor et al. [8] | Heart failure | MLWHF | 17 (1995) | Mean treatment effect -7.1 (-10.5, -3.7) |
| Dallas et al. [57] | Heart failure | Hospitalizations | 26 (4664) | Odds Ratio 0.56 (0.42, 0.75) |
| Tucker et al. [58] | Heart failure with reduced ejection fraction | Left ventricular ejection fraction | 14 (810) | Weighted mean difference 3.79% (2.08, 5.50) for moderate intensity continuous training |
| Lane et al. [59] | Claudication | Pain-free walking distance | 9 (391) | Weighted mean difference 82.1 m (71.7, 92.4) |
| Lane et al. [59] | Claudication | 'Physical summary score' from SF-36 at 6 months | 5 (429) | Weighted mean difference 2.15 (1.26, 3.04) |
| Salman et al. [60] | COPD | Shortness of breath by Chronic Respiratory Disease Questionnaire | 12 (723) | Standardized effect size -0.62 (-0.91, -0.26) |
| Paneroni et al. [61]<sup>b</sup> | Severe COPD | SGRQ | 5 (182) | Weighted mean difference -0.04 (-15.3, -0.8) |
| Baillet et al. [28]<sup>b</sup> | Rheumatoid arthritis | HAQ | 9 (771) | Standardized mean difference 0.24 (0.10, 0.38) |
| Baillet et al. [62]<sup>c</sup> | Rheumatoid arthritis | Joint count | 6 (390) | Weighted mean difference -5.36% (95% CI -9.00, -1.72%) |
| Liang et al. [32] | Cervical radiculopathy | Neck disability index | 5 (514) | Standardized mean difference -1.49 (-2.82, -0.17) |
| Sosa-Reina et al. [34] | Fibromyalgia | Fibromyalgia impact scale | 11 (980) | Standardized mean difference -0.67 (-0.89, -0.45) |
| Sanders et al. [63] | Cognitive impairment | Global cognition | 11 (699) | Hedges' d 0.47 (0.19, 0.74) |
| Jia et al. [64] | Dementia | MMSE score | 13 (673) | Standardized mean difference 1.12 (0.66, 1.59) |
| Rimer et al. [65] | Depression | Depression assessed with different tools | 28 (1101) | Standardized mean difference -0.67 (-0.90, -0.43) |
| Aylett et al. [66] | Anxiety | Anxiety score | 6 (194) | Hedges' d -0.41 (-0.70, -0.12) |
| Sabe et al. [67] | Schizophrenia | PANSS-N | 17 (953) | Standardized mean difference -0.24 (-0.42, -0.06) |
| Sabe et al. [67] | Schizophrenia | PANSS-P | 16 (935) | Standardized mean difference -0.18 (-0.34, -0.02) |

<sup>a</sup> All estimates reported in this table favor exercise groups; effect sizes are as reported by the authors of original meta-analyses.
<sup>b</sup> The meta-analysis includes only aerobic training trials.
<sup>c</sup> The meta-analysis includes only resistance training trials.
**Abbreviations:** MLWHF = Minnesota Living with Heart Failure questionnaire; SF-36 = Short Form health survey; SGRQ = St. George's Respiratory Questionnaire; HAQ = Health Assessment Questionnaire; MMSE = Mini-Mental State Examination; PANSS-N = Negative symptoms – Positive and Negative Symptoms Scale; PANSS-P = Positive symptoms – Positive and Negative Symptoms Scale.

Table 4 provides a summary of findings concerning disease-specific indicators used as outcomes in the meta-analyses. In T2D patients, improved levels of HbA_1c_ percent after exercise therapy reflect improved disease status. In hypertension, reductions in systolic and diastolic blood pressure levels indicate improvement, while a reduction in the need for hospital admissions is seen with coronary heart disease. In heart failure, improvements are gauged by the Minnesota Living with Heart Failure Questionnaire (MLWHFQ) [68, 69], which is one of the most widely used disease-specific HRQoL questionnaires for patients with heart failure. It provides scores for two dimensions, physical and emotional, and a total score. In heart failure with reduced ejection fraction, exercise therapy helps to improve the ejection fraction. In patients with claudication, improvement in disease status is indicated both by longer pain-free walking distance and the physical summary score from the Short Form 36 (SF-36) health survey [70].

In COPD, improvements in shortness of breath are seen by The Chronic Respiratory Disease Questionnaire, the most commonly used disease-specific measurement tool to assess HRQoL in patients with chronic respiratory disease [71, 72]. In patients with severe COPD, improvements have also been reported on St. George’s Respiratory Questionnaire (SGRQ) [73, 74] (Table 4).

The Health Assessment Questionnaire has been developed to measure functional outcomes of patients with arthritis and is a valid self-reported measure of patient functional status over time [75, 76]. It has been widely used in rheumatoid arthritis patients, and studies show that exercise improves this score among such patients. In addition, exercise therapy improves scores on the neck disability index in patients with cervical radiculopathy and on the fibromyalgia impact scale in patients with fibromyalgia (Table 4).

In neurological and psychiatric diseases, exercise therapy improves cognitive outcomes among patients with cognitive impairment, reduces depression in patients with depressive disorders and anxiety in patients with anxiety disorders, and reduces symptoms among patients with schizophrenia (Table 4).

### Effects on health-related quality of life

Usually, when exercise therapy trials have documented general well-being or quality-of-life outcomes, some degree of trend towards improvement is seen. Convincing findings of improvement in HRQoL from meta-analyses are available for knee and hip osteoarthritis, rheumatoid arthritis, fibromyalgia, Parkinson’s disease, schizophrenia, and chronic kidney disease, along with cancer survivors (Table 5) [25, 27, 28, 35, 36, 43, 46, 77-84]. When reporting on quality of life, possible sub-domains that are improved in the absence of improvement in total scores are not formally reported in this review. It should be noted that sub-domains related to physical function are improved in almost all diseases, leading to less physical disability. Improvements in physical function and mood already appear several weeks to months before better HRQoL. To some extent, quality-of life scores overlap with the disease-specific scores of disease progression, and their use varies in specific trials and with specific diseases. HRQoL may see greater improvement if a patient finds exercise therapy enjoyable.

**Table 5.** Effects of exercise therapy on the health-related quality of life benefits, based on selected meta-analyses of randomized controlled trials in the treatment of patients with specific diseases.

| Study | Disease | Outcome measure | N trials<br>(N participants) | Effect size of exercise compared to controls, pooled statistics<br>(95% CI)<sup>a</sup> |
| --- | --- | --- | --- | --- |
| Long et al. [77] | Heart failure | Health-related quality of life | 26 (3833) | Standardized mean difference 0.60<br>(0.36, 0.82) |
| Fransen et al. [25] | Knee osteoarthritis | Quality of life | 13 (1073) | Standardized mean difference 0.28<br>(0.15, 0.40) |
| Goh et al. [27] | Hip or knee osteoarthritis | Quality of life | 33 (2629) | Standardized mean difference 0.21<br>(0.11, 0.31) |
| Baillet et al. [28]<sup>b</sup> | Rheumatoid arthritis | Health-related quality of life | 5 (586) | Standardized mean difference 0.39<br>(0.23, 0.56) |
| Bidonde et al. [35]<sup>b</sup> | Fibromyalgia | Health-related quality of life | 5 (372) | Difference in absolute improvement 8%<br>(3%, 13%) |
| Bidonde et al. [36] | Fibromyalgia | Health-related quality of life | 13 (610) | Difference in absolute improvement 7%<br>(3%, 11%) |
| Dauwan et al. [43] | Schizophrenia | Quality of life | 5 (218) | Hedges' g 0.89<br>(0.22, 1.55) |
| Goodwin et al. [78] | Parkinson's disease | Health-related quality of life limitations | 4 (292) | Standardized mean difference -0.27<br>(-0.51, -0.04) |
| Dauwan et al. [43] | Parkinson's disease | Quality of life | 19 (1739) | Hedges' g 0.31<br>(0.08, 0.54) |
| Dauwan et al. [43] | Multiple sclerosis | Quality of life | 25 (1550) | Hedges' g 0.41<br>(0.24, 0.58) |
| Mishra et al. [79] | Cancer survivors | Health-related quality of life | 11 (826) | Standardized mean difference 0.48<br>(0.16, 0.81) |
| Chen et al. [80] | Advanced-stage cancer | Quality of life | 8 (564) | Standardized mean difference 0.22<br>(0.06, 0.38) |
| Soares Falcetta et al. [81] | Women after treated breast cancer | Quality of life | 24 (1961) | Standardized mean difference 0.45<br>(0.20, 0.69) |
| Singh et al. [46] | Stage II+ breast cancer | Quality of life | 40 (3374) | Standardized mean difference 0.40<br>(0.33, 0.47) |
| Hong et al. [82] | Breast cancer survivors | Quality of life | 18 (1205) | Standardized mean difference 0.35<br>(0.15, 0.54) |
| Larun et al. [83] | Chronic fatigue syndrome | Self-perceived (positive) change in overall health | 4 (489) | Risk Ratio 1.83<br>(1.39, 2.40) |
| Pei et al. [84]<sup>b</sup> | Chronic kidney disease | Quality of life | 7 (562) | Standardized mean difference 8.90<br>(2.48, 15.32) |

<sup>a</sup> All estimates reported in this table favor exercise groups; effect sizes are as reported by the authors of original meta-analyses.
<sup>b</sup> The meta-analysis includes only aerobic training trials.

## Comments

### From evidence to real life

This review shows that exercise therapy has a wide variety of beneficial effects among patients with specific chronic diseases. A systematic review of complications was not a focus of this review, but the meta-analyses of the RCTs did not report an excess of severe complications in exercise groups compared to control groups. However, it should be kept in mind when tailoring exercise therapy interventions for patients that the researchers have considered the main contraindications for exercise in their study inclusion criteria and in the baseline screenings of participants.

This review has not extensively discussed the differences between the effects of different types of training. In many disease conditions, different exercise modalities have rather similar effects on health outcomes, and the key factor in real-life settings may be the patients selecting their preferred type(s) of exercise to achieve long-term therapeutic benefits [85]. However, it should be noted that very specific types of exercise may include some advantages in the treatment of specific diseases, such as dancing in Parkinson’s disease [86]. In addition, a combination of types of training may be beneficial, as endurance and strength training and varying intensities of training may be due to their differing to some extent from one another; additional mechanisms may increase effect sizes, such as combining different types of training in the treatment of non-insulin-dependent diabetes [87].

Since long-term adherence is a general problem in exercise therapy, supervised exercise programs often lead to better results than non-supervised programs [24], although the cost-effectiveness of programs with little or no supervision may be higher. In some patient groups and intervention types, supervision of patient groups at the beginning of the intervention is indicated, which may be more cost-effective than individually supervised interventions. Cost-effectiveness was not a focus of this review. There is a limited amount of good research available on the cost-effectiveness of exercise therapy, but there are data supporting the cost-effectiveness of exercise therapy in the treatment of patients with coronary heart disease, chronic heart failure, intermittent claudication [88], and sub-acute or chronic low back pain [89]. In RCTs, all exercise intervention patients usually have an identical, predetermined training program. In real life, this is not an optimally cost-effective way to tailor exercise therapies.

When individual parameters such as disease stage, physical fitness, motivational factors, exercise preferences, and the infrastructure available for exercising in the nearby everyday environment are considered, the feasibility of exercise therapy and its cost-effectiveness can most likely be improved. In addition, patients need to be followed and personalized support and supervision need to be increased if patients are unable to respond to low-cost exercise recommendations.

When tailoring exercise therapy and possibly selecting objective physical activity monitoring devices, the differences between absolute exercise intensity and exercise intensity relative to fitness level need to be understood [90]. If the monitoring devices available do not permit individual calibration of exercise intensities, perceived exercise intensity levels can be used for guidance. The goals of the therapy and optimal exercise mode may differ by disease condition. To recover from an acute musculoskeletal condition, a short but intensive muscle training program may be the best option. When trying to achieve cardio-metabolic benefits, it should be noted that the primary goal is long-term behavioral change with an influence on long-term disease progression, although benefits such as increased fitness levels and reduced depression can be achieved even during a short intervention. In addition, in real-life clinical work, it is important not only to increase the physical activity level but also to correct other health behaviors: diet, smoking, use of alcohol, and so on. Participation in exercise programs such as supervised exercise at a gym often leads to dietary changes as a co-intervention, and these are generally not carefully controlled in most published trials.

### Methodological considerations and main limitations

There are some limitations in the studies on which the meta-analyses are based. First, many exercise therapy studies have involved fairly low numbers of participants to document uncommon severe complications and, due to the nature of the intervention, they are less often rigorously blinded or placebo-controlled than pharmacological clinical trials. While randomization and intention-to-treat analyses have been carried out in recent RCTs, there appears to be an ongoing problem with concealment of allocation, as in many meta-analyses the effects are not as strong in the sub-group of studies where concealment of allocation has been properly described. This also explains why effects of exercise therapy on mortality did not reach a high enough standard of evidence to be included in the present study [91, 92]. A common problem in exercise therapy studies is the insufficient documentation and analysis of possible co-interventions, such as changes in medication or diet. Also, improvement in medications both in exercise and control groups may reduce the independent effects of exercise on disease progression. Generalizability may be a further problem, as many RCTs include patients who are not representative of the general population of patients with respect to age, gender, exercise motivation, and coexisting diseases. In RCTs, volunteer patients may be more motivated to follow exercise therapy recommendations than the overall patient population in real life, so interviews on motivational factors are important in clinical practice. The limitations of this review include its having only one author and a novel review process design; however, the author has a long track record of clinical experience in sports and exercise medicine and in conducting traditional systematic reviews.

### Future challenges and conclusion

Evidence of many health benefits of exercise therapy for patients with chronic disease has increased substantially over the past decade. Although meta-analyses including at least four RCTs are not available for all diseases and outcomes, there is promising preliminary evidence for the beneficial effects of exercise therapy in some less frequently studied chronic diseases; still, more high-quality studies with longer follow-ups are needed. Recently, new types of meta-analyses have been increasingly favored to advance science. Network meta-analyses using both direct and indirect comparisons of different types of exercise therapies have provided additional information on the effectiveness of the therapies [93, 94], although this approach includes the limitation that indirect evidence with only a small number of direct pairwise comparisons may lead to biased conclusions. In addition, data are accumulating from individual participant meta-analyses, which may help in tailoring individually optimized exercise therapies in the future.

Physicians play an important role in paying attention to the physical activity habits of their patients, in evaluating their risks related to vigorous exercise, and in motivating patients when prescribing exercise to those with chronic disease. Collaboration with an exercise physiologist, physiotherapist, or similar allied professional will be beneficial when determining correct exercise intensities and in tailoring and supervising training programs. It is a challenge to know and improve the local collaborative infrastructure to support prolonged exercise participation. A good start to improve treatment practice is to document in patients with chronic disease whether physical activity is already part of their lives and to encourage them to increase physical activity when indicated, based on the findings of this review.

## Data Availability

All data are provided in the article and the Supplementary file 1.

## Declarations

### Data Availability

All data are provided in the article and the Supplementary file 1.

### Authorship Contributions

All contributions by UMK.

