## Supplementary file 1 for "Summary of the Effects of Exercise Therapy in Non-Communicable Diseases: Clinically Relevant Evidence from Meta-Analyses of Randomized Controlled Trials"

Supplementary file 1 to article:

### Summary of the Effects of Exercise Therapy in Non-Communicable Diseases: Clinically Relevant Evidence from Meta-Analyses of Randomized Controlled Trials

Author: Prof Urho M Kujala, MD, PhD, Faculty of Sport and Health Sciences, P.O. Box 35 (LL), FIN-40014 University of Jyväskylä, Finland. Tel. +358 40 805 3567,

**Supplementary Table 1. Search terms and accumulation of hits in searches* of meta-analyses of exercise therapy research in PubMed and Cochrane Database of Systematic Reviews.**

| Search terms | Hits through Jan 1, 2001 | Hits through Jan 1, 2011 | Hits through Jan 1, 2021 |
|---|---|---|---|
| **PubMed†** | | | |
| Metabolic syndrome | 1 | 8 | 58 |
| Diabetes | 7 | 35 | 291 |
| Hypertension | 9 | 27 | 182 |
| Coronary heart disease | 16 | 31 | 98 |
| Myocardial infarction | 15 | 34 | 95 |
| Heart failure | 10 | 55 | 268 |
| Claudication | 5 | 14 | 52 |
| Stroke | 3 | 25 | 157 |
| Asthma | 3 | 8 | 41 |
| Chronic obstructive pulmonary disease | 6 | 32 | 168 |
| Osteoarthritis | 1 | 15 | 144 |
| Rheumatoid arthritis | 1 | 9 | 26 |
| (Low) back pain | 3 | 27 | 151 |
| Neck pain | 1 | 6 | 50 |
| Fibromyalgia | 1 | 7 | 40 |
| Osteoporosis | 2 | 12 | 55 |
| Depression | 11 | 42 | 311 |
| Anxiety | 4 | 19 | 166 |
| Schizophrenia | 0 | 1 | 35 |
| Dementia | 0 | 16 | 158 |
| Parkinson’s disease | 0 | 3 | 67 |
| Multiple sclerosis | 0 | 5 | 53 |
| Cancer | 2 | 28 | 300 |
| Chronic fatigue syndrome | 0 | 0 | 11 |
| Kidney disease | 0 | 7 | 51 |
| HIV positive individuals | 0 | 6 | 26 |
| **Cochrane Database of Systematic Reviews** | | | |
| Exercise (in title, abstract or as keyword) | | | 673 |

*Searches were carried out between Jan 1 and Jan 30, 2021; note that there is overlap between searches and that, practically speaking, the same meta-analyses can be published in the Cochrane Library and a journal.

†A typical search in PubMed concerning hits through Jan 1, 2021: (((exercise[Title/Abstract]) AND (meta-analysis[Title/Abstract])) AND (disease-specific terms included here[Title/Abstract])) AND (("1900/01/01"[Date - Publication] : "2021/01/01"[Date - Publication]))
